# Integrating Heat-Stable Carbetocin into Routine Maternal Care: Lessons from District-wide Implementation of an AMTSL Strengthening Model in India

**DOI:** 10.64898/2026.08.19.26360867

**Authors:** Harish Kumar, Saurabh Bhargava, Archana Mishra, Naresh Chandra Joshi, Anil Nagedra, Sachin Gupta

## Abstract

Postpartum haemorrhage (PPH) remains the leading direct cause of maternal mortality globally, with a disproportionate burden in low- and middle-income countries. Although prophylactic uterotonics are effective, their impact is often constrained by health system limitations, including unreliable cold-chain storage affecting oxytocin quality. Heat-stable carbetocin (HSC) offers a thermally stable alternative; however, evidence on its large-scale integration into routine public health systems remains limited.

We conducted a district-wide implementation evaluation of an HSC-based Active Management of the Third Stage of Labour (AMTSL) strengthening model across 32 public-sector delivery facilities in Dewas district, Madhya Pradesh, India. Implemented through a phased public–private partnership, the model integrated HSC into routine labour room practice alongside provider capacity building, strengthened documentation, and supportive supervision. A retrospective observational design was used to analyse routinely collected facility-level data from August 2022 to December 2024. Key outcomes included prophylactic uterotonic coverage, timeliness of administration, PPH incidence, and management practices. A total of 48,487 institutional deliveries were recorded during the study period. Documented prophylactic uterotonic coverage was nearly universal (99.9%), with administration within one minute of birth achieved in 99.4% of deliveries. Among deliveries with documented prophylactic uterotonic use, 41,658 (85.9%) received HSC and 6,812 (14.1%) received oxytocin. Overall, 275 PPH cases (0.57%) were documented. Among women receiving HSC prophylaxis, 200 (0.48%) developed PPH, compared with 75 (1.10%) among those receiving oxytocin. These findings are descriptive because prophylactic uterotonic allocation reflected routine programme implementation rather than random assignment. Uterine atony was the leading documented cause of PPH (176/275; 64.0%). Management included tranexamic acid in 239 (86.9%) cases, intravenous fluids in 273 (99.3%), blood transfusion in 36 (13.1%), and referral to a higher-level facility in 84 (30.5%) cases. HSC uptake was significantly higher in First Referral Units than non-FRU facilities (89.3% vs. 81.4%; p<0.001), as was administration within one minute among HSC recipients (100% vs. 98.7%; p<0.001).

District-wide implementation of an HSC-based AMTSL strengthening model achieved high coverage and timely administration of prophylactic uterotonics across public-sector facilities operating at different levels of obstetric capacity. The findings provide real-world implementation evidence supporting the feasibility of integrating HSC into routine government maternity services using existing health-system infrastructure, supervision, and reporting mechanisms. Such embedded implementation approaches may offer a pragmatic pathway for strengthening PPH prevention in settings where reliable maintenance of the oxytocin cold chain remains challenging.

## Introduction

Postpartum haemorrhage (PPH) remains the leading direct cause of maternal mortality globally, accounting for approximately 27% of all maternal deaths worldwide, with the highest burden concentrated in low- and middle-income countries (LMICs), particularly in South Asia and sub-Saharan Africa [1,2]. Despite the availability of effective preventive interventions, most haemorrhage-related deaths occur during the intrapartum or immediate postpartum period, reflecting persistent gaps in the quality and reliability of routine maternity care rather than a lack of clinical knowledge or technologies [1].

India contributes substantially to the global maternal mortality burden. Although the country has reduced its maternal mortality ratio (MMR) by nearly 70% over the past two decades, progress has been uneven across states [3]. According to the latest Sample Registration System (SRS) Special Bulletin on Maternal Mortality in India (2022–2024), India’s maternal mortality ratio (MMR) is estimated at 87 deaths per 100,000 live births. Although substantial progress has been achieved, Madhya Pradesh continues to report one of the highest MMRs in the country at 135 deaths per 100,000 live births, second only to Uttar Pradesh (154), highlighting the continued need for targeted interventions to address preventable causes of maternal mortality, including postpartum haemorrhage [4]. National mortality analyses identify obstetric haemorrhage as a leading direct cause of maternal death in India, with a disproportionate burden in high-burden states such as Madhya Pradesh [5].

The World Health Organization (WHO) and International Federation of Gynecology and Obstetrics (FIGO) recommendations identify timely administration of a prophylactic uterotonic during the third stage of labour as the cornerstone of postpartum haemorrhage prevention and a key component of Active Management of the Third Stage of Labour (AMTSL) [6,7]. Oxytocin, the first-line uterotonic recommended by the World Health Organization (WHO), is widely used in India’s public health system. However, oxytocin is highly thermolabile and requires uninterrupted cold-chain storage at 2–8°C. Evidence from India and other LMICs has documented frequent cold-chain disruptions, improper storage and transport, and variable medicine quality under routine program conditions particularly in peripheral and high-volume public-sector facilities compromising the effectiveness of AMTSL in practice [8–10].

The WHO recommend consideration of heat-stable carbetocin (HSC) for prevention of PPH in settings where the quality of oxytocin cannot be reliably assured and where its cost is comparable to other effective uterotonics [11]. HSC is a long-acting oxytocin analogue with superior thermal stability, retaining potency at temperatures up to 30–40°C, making it operationally well suited to LMIC health systems [12]. A large WHO-led multi-country randomised controlled trial demonstrated that HSC is non-inferior to oxytocin for PPH prevention [13]. In response, Injectable HSC (100 μg/mL) was subsequently included in the WHO Model List of Essential Medicines, and in 2020, the Drug Controller General of India (DCGI) approved its use for postpartum haemorrhage prevention, creating an opportunity to strengthen uterotonic coverage within India’s public health system [14, 15].

Despite strong clinical evidence and normative guidance, global analyses increasingly emphasise that implementation gaps rather than lack of effective interventions constitute the principal barrier to reducing PPH-related mortality [16]. Large-scale, real-world evidence on the integration of heat-stable uterotonics into routine public-sector maternity services, particularly in high-burden settings, remains limited.

In Madhya Pradesh, the USAID-supported SAMVEG project, implemented by IPE Global in partnership with the National Health Mission (NHM) and Ferring Pharmaceuticals, introduced heat-stable carbetocin as part of a broader AMTSL strengthening initiative within the public health system. The intervention was implemented through a public–private partnership (PPP) model that integrated HSC into routine labour room practice alongside provider capacity building, strengthened documentation, supportive supervision, and alignment with government drug supply systems.

Public-sector maternity services in India are delivered through a tiered referral network comprising Sub-Centres (SCs), Primary Health Centres (PHCs), Community Health Centres (CHCs), District Hospitals (DHs), and higher referral hospitals. These facilities provide progressively higher levels of maternal and newborn care, with designated First Referral Units (FRUs) equipped to provide Comprehensive Emergency Obstetric and Neonatal Care (CEmONC), including caesarean section, blood transfusion, and management of obstetric emergencies, in accordance with the Indian Public Health Standards (IPHS) (17). Women with complications identified at lower-level facilities are referred to FRUs for definitive care. This existing referral architecture provided the platform for implementation and subsequent district-wide scale-up of the intervention.

An initial pilot implementation across 15 public health facilities in Dewas district, covering 18,497 institutional deliveries, demonstrated the feasibility and safety of integrating HSC into routine maternity care and has been reported previously [18]. However, the pilot was limited in scope and not designed to assess system performance at scale. Building on these findings, the National Health Mission expanded the HSC model across all 32 public-sector delivery facilities spanning the district referral network, covering approximately 48,000 institutional deliveries between August 2022 and December 2024.

This district-wide implementation provides a unique opportunity to evaluate the feasibility, consistency, and system-level performance of integrating heat-stable carbetocin into public-sector maternity services at scale. This study examines program coverage, facility readiness, provider adherence to AMTSL practices, and trends in PPH-related outcomes under routine service conditions, generating policy-relevant evidence to inform decisions on broader adoption of heat-stable uterotonics in high-burden LMIC settings.

## Materials and Methods

### Description of the Intervention

The HSC-based AMTSL strengthening model was implemented through a phased approach, starting from situational assessment, co-design to pilot implementation in selected health facilities and subsequently district-wide scale-up covering all health facilities. The intervention was co-created through a public–private partnership involving the National Health Mission (NHM), Madhya Pradesh; the USAID-supported SAMVEG project implemented by IPE Global; and Ferring Pharmaceuticals. The intervention comprised five interrelated components: (i) introduction of HSC into routine labour room practice; (ii) competency-based training of healthcare providers on AMTSL and appropriate HSC use; (iii) strengthening of labour room documentation and routine monitoring systems; (iv) supportive supervision and mentoring; and (v) integration with existing government procurement, supply chain, and referral systems through a public–private partnership. The overall implementation pathway is illustrated in Fig 1.

**Fig 1:**
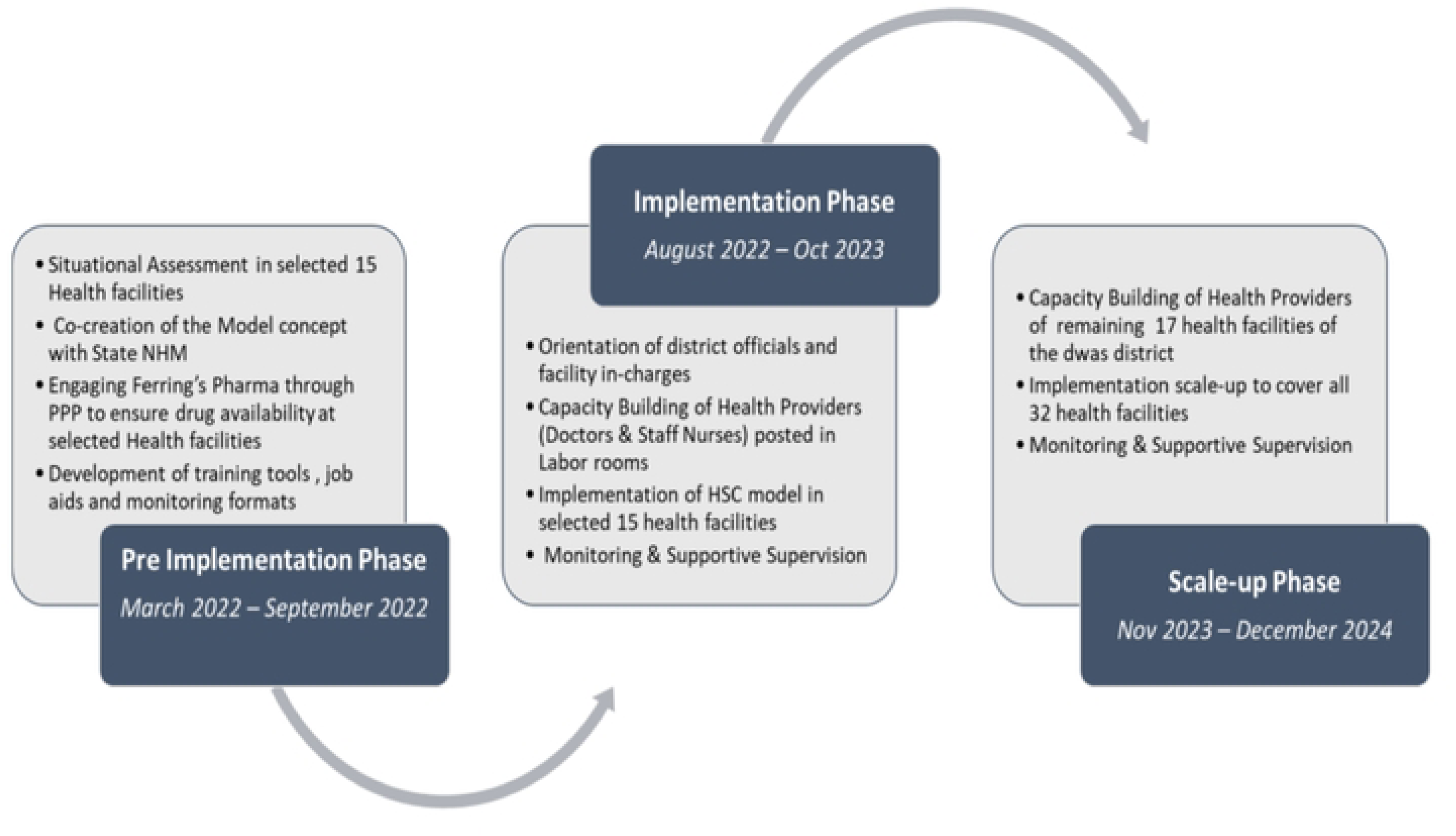
Phased implementation of the heat-stable carbetocin–based AMTSL strengthening model in Dewas district, Madhya Pradesh.

#### Pre-implementation phase (March 2022 – September 2022)

The pre-implementation phase focused on assessment of existing system gaps and co-creating a contextually appropriate intervention model. A structured situational assessment was conducted in 15 selected public health facilities to examine routine AMTSL practices, uterotonic availability and use, cold-chain functionality, provider knowledge and adherence to national guidelines, labour room documentation, and immediate postpartum monitoring.

The assessment identified key constraints affecting reliable prevention of postpartum haemorrhage, including inconsistent documentation of uterotonic administration and timing, variable adherence to AMTSL protocols across cadres, unreliable cold-chain infrastructure particularly in non-FRU facilities and limited routine monitoring during the immediate postpartum period. Although oxytocin was widely available, concerns were noted regarding its quality at the point of care due to storage and cold-chain interruptions. These findings highlighted that implementation and system-readiness gaps, rather than lack of clinical guidance, were the primary barriers to effective PPH prevention.

Using these insights, the intervention model was co-created with the State NHM using human-centred design approach, emphasising alignment with frontline workflows, facility constraints, and existing health-system processes. Iterative discussions with state and district program managers, clinicians, and nursing staff informed the refinement of model components, implementation sequencing, and documentation tools. During this phase, Ferring Pharmaceuticals was engaged through a public–private partnership to support uninterrupted supply of heat-stable carbetocin at demonstration health facilities using the state’s drug distribution system.

A Technical Advisory Group (TAG) was constituted for development of training materials, job aids and monitoring formats in alignment with national AMTSL guidelines, focusing on the appropriate use of HSC, including contraindications to its administration to support subsequent implementation. Local language adaptation was done after obtaining State Government approvals for the training materials. Capacity-building activities were implemented through the state government’s established training infrastructure and logistics systems, with sessions facilitated by government-recognised trainers.

#### Implementation phase (August 2022 – October 2023)

The implementation phase involved pilot deployment of the model in the same 15 public health facilities. This phase began with orientation of district health officials and facility in-charges to ensure administrative alignment and ownership.

Capacity-building activities were conducted for all doctors and staff nurses posted in labour rooms of 15 selected health facilities, focusing on correct AMTSL practices, appropriate timing and contraindications for uterotonic administration, and avoidance of misuse of uterotonics for labour induction or augmentation. HSC was introduced into routine labour room practice as the prophylactic drug for PPH prevention.

Implementation was supported through monthly monitoring and supportive supervision by IPE Global project staff, using the monitoring formats developed during the pre-implementation phase. Program data generated during this phase were used to assess feasibility, provider acceptance, and safety of the model under routine service conditions and to generate evidence to inform decisions on scale-up.

#### Scale-up phase (November 2023 – December 2024)

Following review of pilot-phase findings and approval by the National Health Mission, the intervention was scaled up to all 32 public-sector delivery facilities in Dewas district. Capacity-building activities were extended to health providers in the remaining 17 facilities, and HSC was integrated into routine AMTSL practice across all delivery points. Scale-up was implemented under routine service delivery conditions, without additional staffing or parallel reporting systems. Ongoing monitoring and supportive supervision were conducted through existing district mechanisms to reinforce adherence to AMTSL practices, ensure appropriate use of HSC, and maintain documentation quality across facilities with varying levels of obstetric capacity.

### Study Design

We conducted a district-wide implementation evaluation of HSC based AMTSL strengthening model implemented across public health facilities in Dewas district, Madhya Pradesh, India. The study adopted a retrospective observational design, analysing routinely collected facility-level program data generated between August 2022 and December 2024 under real-world service delivery conditions.

The evaluation period spanned both the pilot implementation phase and the subsequent district-wide scale-up. The initial 15 facilities that included in the implementation phase contributed data from August 2022 onward, whereas the additional 17 facilities that joined during the scale-up phase contributed data from November 2023 onward, corresponding to the timing of model expansion.

The evaluation was designed to assess implementation under routine programmatic conditions rather than to compare the clinical effectiveness of HSC and oxytocin. Accordingly, the analysis focused on programme coverage, implementation fidelity, adoption across facility levels, and routine maternal health outcomes.

### Study Settings and Population

The study included all 32 public health facilities in Dewas district, comprising one District Hospital, seven Community Health Centres, 22 Primary Health Centres, and two Sub-Centres. Of these, three facilities were designated as FRUs providing CEmONC services, while the remaining 29 facilities functioned as non-FRU delivery points providing routine maternity services. The inclusion of facilities across multiple levels of the district referral network enabled evaluation of implementation under varying levels of obstetric capacity, referral capability, and health system readiness. All women delivering in these facilities during the implementation period were included.

### Data Collection

Rather than introducing parallel reporting systems, the intervention embedded structured documentation within established labour room registers and delivery records. Hence, data for this evaluation were derived from routinely maintained facility-level records and district monitoring systems. During the pre-implementation phase, existing labour room and delivery registers were reviewed and additional data fields were incorporated into existing recording formats to strengthen routine monitoring and improve fidelity of implementation.

Revised formats included fields capturing the type of prophylactic uterotonic administered (HSC or oxytocin), timing of uterotonic administration relative to birth, documentation of PPH diagnosis, and key management measures such as administration of additional uterotonics, tranexamic acid use, blood transfusion, and referral. Facility staff were oriented on accurate and complete recording of these fields as part of the capacity-building activities conducted during the implementation and scale-up phases.

Throughout the study period, data were recorded by routine service providers in labour room registers and subsequently compiled into monthly facility reports and shared with the district nodal person based at the NHM for district-level compilation and monitoring. PPH cases were identified through facility-level PPH line listings maintained as part of routine obstetric monitoring processes. The project staff provided monthly supportive supervisory visits included review of documentation completeness and internal consistency of recorded indicators to support data quality.

### Outcome Measures

Outcome measures included delivery characteristics, AMTSL performance indicators, and postpartum haemorrhage related outcomes. Delivery indicators comprised total institutional deliveries, vaginal deliveries, and caesarean sections recorded during the study period. AMTSL performance was assessed through prophylactic uterotonic administration following delivery, type of uterotonic used (HSC or oxytocin), and timeliness of administration within one minute of birth. Postpartum haemorrhage indicators included the number and proportion of documented PPH cases, presumed causes and management measures such as use of additional uterotonics, tranexamic acid, blood transfusion, and referral. Maternal deaths attributed to obstetric haemorrhage were also recorded. Indicators were summarised at district level and stratified by facility designation (FRU versus non-FRU) where relevant.

### Data Analysis

We performed facility-level analysis using Microsoft Excel (Office 365) and IBM SPSS Statistics version 26, and the results are reported in aggregated form in terms of frequencies and proportions. Descriptive analyses focused on implementation indicators including intervention coverage, timeliness of uterotonic administration, facility-level adoption, and postpartum haemorrhage outcomes. Comparisons between FRU and non-FRU facilities were undertaken to examine implementation performance across different levels of the district health system.

Additionally, the association between HSC use for PPH prevention and level of health facility was calculated and compared for statistical significance using Pearson’s chi-square test.

### Ethical Considerations

We received administrative approval from the NHM, Madhya Pradesh before conducting this analysis. Additionally, this study was reviewed and approved by Institutional Review Board (IRB) of the International Institute of Health Management Research (IIHMR), New Delhi (IRB No. IIHMR D/SRB/2/2022). The analysis used routinely collected programme data and did not involve prospective recruitment of participants. The dataset was accessed for research purposes on 20 June 2025. The authors did not have access to information that could identify individual participants during or after data collection.

## Results

The HSC based AMTSL strengthening model was implemented in a phased manner between August 2022 and December 2024, beginning with a pilot implementation phase followed by district-wide scale-up. During this period, a total of 142 healthcare providers were trained, including 88 during the initial implementation phase and 54 during the scale-up phase. Staff nurses constituted 81% of the total trained personnel, reflecting their primary role in intrapartum and immediate postpartum care in labour rooms. (Table 1).

**Table 1.** Capacity building of healthcare providers during implementation and scale-up of the HSC model.

| Cadre | Implementation Phase n (%) | Scale-up Phase n (%) | Total n (%) |
| --- | --- | --- | --- |
| Doctors | 18 (20.5) | 9 (16.7) | 27 (19.0) |
| Staff Nurses | 70 (79.5) | 45 (83.3) | 115 (81.0) |
| **Total** | **88 (100.0)** | **54 (100.0)** | **142 (100.0)** |

During the study period, 50,029 deliveries were reported across participating facilities. Of these, 1,542 (3.1%) women delivered before reaching a health facility and were managed as outside (transit) deliveries. After excluding these cases, 48,487 institutional deliveries were included in the analysis. Nearly half of all institutional deliveries (24,079; 49.7%) occurred at the District Hospital, followed by Community Health Centres (12,175; 25.1%), Primary Health Centres (11,814; 24.4%), and Sub-Centres (419; 0.9%). Overall, 41,255 (85.1%) deliveries were normal vaginal deliveries, while 7,232 (14.9%) were lower segment caesarean sections (LSCS) conducted at FRU facilities, consistent with referral norms and availability of surgical obstetric services (Table 2).

**Table 2.** Coverage and timely administration of prophylactic uterotonics for postpartum haemorrhage prevention by facility level and mode of delivery. † *Percentages for administration within 1 minute of birth were calculated using the number of women receiving the respective prophylactic uterotonic (HSC or oxytocin) during the corresponding mode of delivery as the denominator*.

| Indicator | DH<br>(n=1) | CHC<br>(n=7) | PHC<br>(n=22) | SC<br>(n=2) | Total<br>(n=32) |
| --- | --- | --- | --- | --- | --- |
| Total deliveries reported during the study period | 24,232 | 13,057 | 12,313 | 427 | 50,029 |
| Outside (transit) deliveries managed | 153 | 882 | 499 | 8 | 1,542 |
| Institutional deliveries included in analysis | 24,079 (49.7%) | 12,175 (25.1%) | 11,814 (24.4%) | 419 (0.9%) | 48,487 |
| Prophylactic HSC administered | 22,037 (91.5%) | 9,725 (79.9%) | 9,528 (80.7%) | 368 (87.8%) | 41,658 (85.9%) |
| Prophylactic oxytocin administered | 2,042 (8.5%) | 2,409 (19.8%) | 2,310 (19.6%) | 51 (12.2%) | 6,812 (14.1%) |
| **Normal Vaginal Deliveries (NVD)** | 16,849 (70%) | 12,173 (100%) | 11,814 (100%) | 419 (100%) | 41,255 (85.1%) |
| HSC administered in NVD | 15,364 (91.2%) | 9,723 (79.9%) | 9,528 (80.7%) | 368 (87.8%) | 34,983 (84.8%) |
| HSC administered within 1 minute of birth<sup>†</sup> | 15,358 (100%) | 9,714 (99.9%) | 9,340 (98.0%) | 353 (95.9%) | 34,765 (99.4%) |
| Oxytocin administered | 1,485 (8.8%) | 2,409 (19.8%) | 2,310 (19.6%) | 51 (12.2%) | 6,255 (15.2%) |
| Oxytocin administered within 1 minute of birth<sup>†</sup> | 1,485 (100.0%) | 2,391 (99.3%) | 2,249 (97.4%) | 51 (100.0%) | 6,176 (98.8%) |
| **Lower Segment Caesarean Section (LSCS) Deliveries** | 7,230 (30.0%) | 2 (<0.1%) | - | - | 7232 (14.9%) |
| HSC administered in LSCS | 6673 (92.3%) | 2 (100%) | - | - | 6675 (92.3%) |
| HSC administered within 1 minute of birth<sup>†</sup> | 6673 (100%) | 2 (100%) | - | - | 6675 (100%) |
| Oxytocin administered in LSCS | 557 (7.7%) | 0 | - | - | 557 (7.7%) |
| Oxytocin administered within 1 minute of birth<sup>†</sup> | 557 (100%) | 0 | - | - | 557 (100%) |

Coverage of prophylactic uterotonics for prevention of PPH was found nearly universal across all facility levels. HSC was the predominant uterotonic used during the implementation period, accounting for 41,658 (85.9%) prophylactic administrations, while oxytocin was used in 6,812 (14.1%) deliveries. Uptake of HSC exceeded 79% across all facility levels and was highest at the District Hospital (91.5%). Among normal vaginal deliveries receiving HSC prophylaxis, administration within one minute of birth was achieved in 99.4% of deliveries. Similarly, among women receiving oxytocin prophylaxis, timely administration within one minute was achieved in 98.8% of deliveries. For caesarean deliveries, HSC was administered in 92.3% of women, while oxytocin was administered in 7.7% of women. Administration within one minute of birth was achieved in all cases receiving either uterotonic. No instances of inappropriate use of HSC for labour induction, augmentation, or therapeutic management of postpartum haemorrhage were documented (Table 2).

Overall, 275 of 48,487 institutional deliveries (0.57%) were complicated by PPH during the study period (Table 3). Community Health Centres accounted for the largest proportion of documented PPH cases (184/275; 66.9%), followed by Primary Health Centres (52/275; 18.9%), the District Hospital (35/275; 12.7%), and Sub-Centres (4/275; 1.5%).

**Table 3.** Postpartum haemorrhage (PPH) incidence among women receiving prophylactic uterotonics by facility level.

| Indicator | DH<br>(n=24,079) | CHC<br>(n=12,175) | PHC<br>(n=11,814) | SC<br>(n=419) | Total<br>(n=48,487) |
| --- | --- | --- | --- | --- | --- |
| **Women receiving Prophylactic HSC** | 22,037 (91.5%) | 9,725 (79.9%) | 9,528 (80.7%) | 368 (87.8%) | 41,658 (85.9%) |
| PPH cases following HSC prophylaxis | 30 (0.14%) | 129 (1.33%) | 37 (0.39%) | 4 (1.09%) | 200 (0.48%) |
| **Women receiving Prophylactic Oxytocin** | 2,042 (8.5%) | 2,409 (19.8%) | 2,310 (19.6%) | 51 (12.2%) | 6,812 (14.1%) |
| PPH cases following Oxytocin prophylaxis | 5 (0.24%) | 55 (2.28%) | 15 (0.65%) | 0 (0.0%) | 75 (1.10%) |
| **Total PPH cases** | 35 (0.15%) | 184 (1.51%) | 52 (0.44%) | 4 (0.95%) | 275 (0.57%) |

Among the 41,658 women who received prophylactic HSC as part of routine programme implementation, 200 (0.48%) developed postpartum haemorrhage (PPH). Across facility levels, the incidence of PPH following HSC prophylaxis ranged from 0.14% at the District Hospital to 1.33% at Community Health Centres. Among the 6,812 women who received prophylactic oxytocin, 75 (1.10%) developed PPH. These findings are presented descriptively, as allocation of prophylactic uterotonics reflected routine programme implementation rather than random assignment (Table 3).

Among the 275 women with documented PPH, uterine atony was the leading cause of PPH, accounting for 176 (64.0%) cases, followed by genital tract tears/lacerations (58; 21.1%), retained placental tissue (24; 8.7%), thrombin disorders (1; 0.4%), and other causes (16; 5.8%) (Table 4).

**Table 4.** Clinical characteristics, management and outcomes of postpartum haemorrhage cases by facility level (n=275)

| Indicator | DH (n=35) | CHC (n=184) | PHC (n=52) | SC (n=4) | Total (n=275) |
| --- | --- | --- | --- | --- | --- |
| **Cause of postpartum haemorrhage** |  |  |  |  |  |
| Atonic uterus | 34 (97.1%) | 113 (61.4%) | 25 (48.1%) | 4 (100.0%) | 176 (64.0%) |
| Tear/laceration | 0 (0.0%) | 40 (21.7%) | 18 (34.6%) | 0 (0.0%) | 58 (21.1%) |
| Retained placental tissue | 0 (0.0%) | 20 (10.9%) | 4 (7.7%) | 0 (0.0%) | 24 (8.7%) |
| Thrombin disorder | 0 (0.0%) | 1 (0.5%) | 0 (0.0%) | 0 (0.0%) | 1 (0.4%) |
| Other causes | 1 (2.9%) | 10 (5.4%) | 5 (9.6%) | 0 (0.0%) | 16 (5.8%) |
| **Management of postpartum haemorrhage** |  |  |  |  |  |
| Tranexamic acid administered | 31 (88.6%) | 160 (87.0%) | 45 (86.5%) | 3 (75.0%) | 239 (86.9%) |
| Intravenous fluids administered | 35 (100.0%) | 184 (100.0%) | 50 (96.2%) | 4 (100.0%) | 273 (99.3%) |
| Blood transfusion given | 10 (28.6%) | 20 (10.9%) | 6 (11.5%) | 0 (0.0%) | 36 (13.1%) |
| **Outcome of postpartum haemorrhage** |  |  |  |  |  |
| Referred | 7 (20.0%) | 59 (32.1%) | 18 (34.6%) | 0 (0.0%) | 84 (30.5%) |
| Discharged | 26 (74.3%) | 125 (67.9%) | 33 (63.5%) | 4 (100.0%) | 188 (68.4%) |
| Maternal deaths | 2 (5.7%) | 0 (0.0%) | 1 (1.9%) | 0 (0.0%) | 3 (1.1%) |

Management of PPH was found consistent with recommended clinical protocols. 239 (86.9%) of PPH cases received tranexamic acid, 273 (99.3%) received intravenous fluids, and 36 (13.1%) required blood transfusion. Referral to a higher-level facility was documented in 84 (30.5%) women, while 188 (68.4%) were discharged following management. Three maternal deaths were reported among women who developed PPH. Among the 275 women with documented PPH, partograph data were available for 273 cases, of whom 260 (95.3%) had documented partograph use during labour. Early initiation of breastfeeding was reported in approximately 97% of institutional deliveries. No surgical intervention for management of PPH, other than repair of genital tract tears or lacerations, was documented during the study period (Table 4).

The Pearson’s chi-square test was also employed to assess the association between the use of HSC for PPH prevention and the type of health facility. The results indicate a significant association, with notably higher utilization of HSC for preventing PPH observed in FRU-level facilities compared to non-FRU facilities (p < 0.001). Additionally, the administration of HSC within one minute of delivery was also significantly more frequent in FRU facilities compared to non-FRU facilities (p < 0.001) [Table 5].

**Table 5.** Association between facility type and uptake of heat-stable carbetocin for postpartum haemorrhage prevention.

| Indicator | FRU | Non-FRU | Total | P Value |
| --- | --- | --- | --- | --- |
| 1. Uptake of prophylactic HSC |  |  |  |  |
| HSC administered | 24,922 (89.3%) | 16,736 (81.4%) | 41,658 (85.9%) | P<0.001 |
| HSC not administered | 3,001 (10.7%) | 3,828 (18.6%) | 6,829 (14.1%) |  |
| Total institutional deliveries | 27,923 | 20,564 | 48,487 |  |
| 2. Timeliness of HSC administration |  |  |  |  |
| HSC administered within 1 minute | 24,916 (100.0%) | 16,524 (98.7%) | 41,440 (99.5%) | P<0.001 |
| HSC administered after 1 minute | 6 (0.02%) | 212 (1.3%) | 218 (0.52%) |  |
| Total HSC administrations | 24,922 | 16,736 | 41,658 |  |

## Discussion

This study presents findings from the district-wide implementation of a HSC based AMTSL strengthening model across public sector delivery facilities in Dewas district, Madhya Pradesh. Building on the previously reported pilot implementation conducted in 15 facilities [18], the present analysis demonstrates that the intervention could be successfully expanded to district scale while maintaining high adherence to recommended AMTSL practices.

Across nearly 48,500 institutional deliveries, prophylactic uterotonic use was almost universal (99.9%), with administration within one minute of delivery achieved in 99.4% of cases. Heat-stable carbetocin was used in 86% of deliveries, indicating substantial uptake within routine service delivery. Importantly, no instances of inappropriate use of carbetocin for labour induction or augmentation were documented, suggesting adherence to recommended clinical indications. High compliance with key process indicators particularly near-universal uterotonic coverage and its timely administration suggests strengthening of routine labour room practices under the intervention. Among documented PPH cases, high recorded use of partograph for intrapartum monitoring (95.3%) further indicates adherence to recommended clinical monitoring practices during labour.

The observed PPH incidence in this study was 0.6%, with uterine atony as the leading cause. This is consistent with reported epidemiological patterns in LMIC settings, where uterine atony accounts for the majority of postpartum haemorrhage cases and obstetric haemorrhage remains a leading contributor to maternal mortality [1,5]. In addition, high adherence to recommended PPH management practices including use of uterotonics, tranexamic acid, and appropriate referral suggests improved readiness of facilities to manage obstetric complications within routine service delivery conditions.

The findings of this study are consistent with a substantial body of global evidence demonstrating the effectiveness of uterotonics in preventing postpartum haemorrhage. The WHO-led CHAMPION trial established that heat-stable carbetocin is non-inferior to oxytocin for prevention of PPH following vaginal birth across diverse settings, including several LMICs [13]. Beyond non-inferiority, carbetocin has pharmacological advantages, including a longer half-life and sustained uterotonic effect, which reduces the need for additional uterotonic dosing in the immediate postpartum period [19]. Evidence from systematic reviews and network meta-analyses further supports the effectiveness of carbetocin in preventing postpartum haemorrhage [19,20]. WHO therefore recommends consideration of heat-stable carbetocin in settings where the quality of oxytocin cannot be reliably assured and where its cost is comparable to other effective uterotonics, highlighting its programmatic relevance for health care systems in LMICs [11]. This recommendation reflects not only its clinical effectiveness but also its operational advantages. Unlike oxytocin, which requires uninterrupted cold-chain storage, heat-stable carbetocin maintains potency at higher ambient temperatures, making it particularly suitable for resource-constrained settings.

A critical challenge in LMIC settings has been the variability in oxytocin quality due to suboptimal storage, transport, and regulatory oversight. Multiple studies from India and other LMICs have documented degradation of oxytocin under suboptimal storage conditions, potentially compromising its effectiveness at the point of care [8–10]. Facility-based audits and simulated client studies in India have further highlighted gaps in storage practices and supply chain management [9]. These findings are consistent with broader global analyses indicating variability in uterotonic quality across resource-constrained settings [8]. In this context, heat-stable carbetocin offers a clear programmatic advantage by eliminating dependence on cold-chain systems and ensuring consistent drug potency.

While the clinical efficacy of HSC has been established through randomized controlled trials, evidence describing its implementation within routine public health systems remains limited. The present study extends the existing evidence by demonstrating that HSC can be successfully integrated into routine government maternity services at district scale while maintaining high programme coverage, near-universal timely administration, and adherence to recommended AMTSL practices. By generating implementation evidence under real-world programmatic conditions, this study addresses an important gap between clinical efficacy and health system adoption. This is consistent with emerging implementation evidence on HSC from low-resource settings, which highlights the importance of integrating the intervention within existing health-system structures and delivery processes [21].

Effective prevention of postpartum haemorrhage depends not only on the availability of efficacious uterotonics but also on health-system factors such as provider competence, adherence to clinical protocols, supportive supervision, and reliable documentation systems. Implementation research emphasises the importance of understanding and strengthening these contextual and system-level factors when translating evidence-based interventions into routine practice [22]. Similarly, evidence on high-quality health systems highlights the importance of competent care, reliable processes, and system capacity in achieving better health outcomes [23]. The high levels of adherence observed in this study, including timely uterotonic administration and appropriate clinical management of PPH, suggest that integrating HSC within a structured implementation model can support these critical components of service delivery.

Evidence from India has documented gaps and variability in adherence to evidence-based intrapartum care practices, including essential practices around childbirth and management of obstetric complications [24,25]. In contrast, the findings from this study indicate that high adherence to recommended practices can be achieved across high-volume public-sector facilities when clinical interventions are combined with structured capacity building and supportive supervision. This is consistent with findings from large-scale quality-improvement initiatives in India, including the WHO Safe Childbirth Checklist programme and the LaQshya initiative, which sought to improve provider practices through structured system-level interventions [26,27].

The present study extends beyond these earlier efforts by demonstrating that a heat-stable uterotonic can be integrated into routine care across multiple levels of the health system at district scale. An important distinguishing feature of this implementation model was its deliberate integration within the existing public health system rather than establishing parallel project-specific delivery mechanisms. The intervention leveraged existing government labour-room infrastructure, human resources, referral pathways, procurement systems, and supervisory mechanisms, thereby embedding HSC within routine maternal health services. This approach facilitated implementation across facilities with different levels of obstetric capacity and supported transition from pilot implementation to district-wide scale-up. The subsequent decision by the Government of Madhya Pradesh to initiate statewide procurement of HSC further demonstrates the potential of embedded implementation approaches to contribute to translation of implementation evidence into health policy and programme adoption.

To our knowledge, this is among the first large-scale implementation evaluations of heat-stable carbetocin within a government health system in India. The findings suggest that combining introduction of a heat-stable uterotonic with structured health-system strengthening while working within existing system capacities can address both clinical and operational considerations in postpartum haemorrhage prevention. This approach may also have contributed to the successful transition from pilot implementation to district-wide scale-up and subsequent policy adoption at the state level.

These findings are further supported by comparison with our earlier pilot study conducted in Dewas district, which showed consistency in key performance indicators, including near-universal uterotonic coverage and high compliance with timely administration following birth [18]. However, the present study extends this evidence substantially by demonstrating that these implementation outcomes can be sustained at scale across facilities with varying levels of obstetric capacity within a routine public health system.

At the same time, the persistence of relatively lower uptake of HSC in non-FRU facilities suggests that differences in facility capacity and implementation readiness may influence adoption patterns across levels of care. These findings highlight the importance of continued supportive supervision and targeted capacity strengthening to ensure equitable adoption across all levels of the health system.

### Strengths and Limitations

This study has several strengths. It represents one of the largest real-world implementation evaluations of heat-stable carbetocin in India, covering a large number of deliveries across facilities with varying levels of obstetric capacity. The use of routinely collected programmatic data enhances the relevance of findings for policy and programmatic decision-making. The phased implementation from pilot to district-wide scale-up provides valuable insights into scalability and health system integration.

However, certain limitations should be acknowledged. The study employed a retrospective observational design without a comparison group, limiting the ability to establish causal relationships between the intervention and observed outcomes. The analysis relied on routinely recorded facility data, which may be subject to reporting inaccuracies or incomplete documentation. Because allocation of prophylactic uterotonics reflected routine programme implementation rather than random assignment, descriptive comparisons between women receiving HSC and oxytocin should not be interpreted as evidence of comparative effectiveness. Additionally, PPH incidence was based on clinical estimation rather than objective measurement of blood loss, which may result in underestimation.

### Policy and Program Implications

The findings from this study have important implications for maternal health policy and program design in India and other LMICs. The demonstration of high coverage and adherence to AMTSL practices at district scale suggests that HSC can be feasibly integrated into routine public health systems when supported by structured implementation approaches, including capacity building, supportive supervision, and strengthened monitoring systems. This is particularly relevant for settings where maintaining consistent quality of oxytocin and cold-chain infrastructure remains a persistent challenge.

Affordability and supply considerations are important determinants of large-scale adoption of newer uterotonics. Ferring Pharmaceuticals has committed to making heat-stable carbetocin available to the public sector in low- and lower-middle-income countries at a subsidised price of US$0.31 ±10% per 100 μg ampoule, on an Ex Works-basis, excluding logistics costs [28]. The current UNFPA Global Supply Catalogue lists heat-stable carbetocin at approximately US$0.49 per 100 μg ampoule, providing a contemporary reference point for international procurement [29]. Together, these pricing mechanisms may help improve the affordability and availability of heat-stable carbetocin for public-sector programmes in resource-constrained settings.

Furthermore, implementation evidence generated through the pilot and district-wide scale-up phases in Dewas district has informed policy discussions at the state level. The Government of Madhya Pradesh has initiated state-wide procurement of heat-stable carbetocin for postpartum haemorrhage prevention in public-sector facilities, reflecting growing confidence in its operational feasibility and programmatic value. This transition from implementation to policy adoption highlights the potential of embedded implementation research to directly inform evidence-based decision-making within health systems. The model demonstrated in this study may therefore provide a practical pathway for other states in India and similar LMIC settings seeking to strengthen postpartum haemorrhage prevention strategies through integration of heat-stable uterotonics.

## Conclusion

The district-wide implementation of a heat-stable carbetocin-based AMTSL strengthening model demonstrates that high coverage and timely administration of prophylactic uterotonics can be achieved within routine public health systems. The findings provide real-world implementation evidence supporting the feasibility of integrating heat-stable carbetocin into routine maternal health services and its potential for scale-up across facilities with varying levels of obstetric capacity. Combining a heat-stable uterotonic with structured capacity building, supportive supervision, and strengthened routine monitoring offers a pragmatic approach to strengthening postpartum haemorrhage prevention in resource-constrained settings.

## Data Availability

The datasets generated and or analysed during the current study are not publicly available because they are derived from routine government health facility records and are subject to government data-access and confidentiality requirements. Data may be made available in aggregated form, where permissible, upon reasonable request to the corresponding author and subject to approval from the relevant government authorities.

## Acknowledgments

The authors acknowledge the support of the National Health Mission, Government of Madhya Pradesh, and the Dewas district health authorities for facilitating implementation of the HSC-based AMTSL strengthening model. We also acknowledge the healthcare providers and program teams across the participating public-sector facilities for their contribution to implementation, routine documentation, and data collection. The authors thank the SAMVEG project team and Ferring Pharmaceuticals Safe Birth Project for their support and cooperation.

